# Statistical Analysis Plan for the Bedside Ultrasound Conducted in Kids with distal upper Limb fractures in the Emergency Department (BUCKLED) trial

**DOI:** 10.1101/2022.06.09.22276178

**Authors:** Philip M Jones, Gerben Keijzers, Joshua Byrnes, Robert S Ware, Peter J Snelling

## Abstract

**Background:** The Bedside Ultrasound Conducted in Kids with distal upper Limb fractures in the Emergency Department (BUCKLED) trial is a multicentre, randomised, open-label, non-inferiority study comparing point-of-care ultrasound to x-ray as the initial imaging modality in children with clinically non-angulated distal forearm injuries.

**Objective:** The purpose of this document is to minimise bias by defining and making publicly available our analysis approach prior to completing the study and reviewing or analysing the trial data.

**Methods:** We developed a statistical analysis plan (SAP) for reporting and analysing the BUCKLED trial including our approach to expected protocol deviations, withdrawals, missing data, and loss to follow up. Study methods are described using a previously published study protocol. Broad statistical analysis principles are outlined. Primary and secondary trial outcomes are described, along with appropriate methods for statistical comparison.

**Results:** This report pre-specifies the SAP for the primary outcome of upper limb function at 4 weeks, using an intention-to-treat analysis with a hypothesis of non-inferiority. Results of a per-protocol analysis will also be reported. Subgroup analyses will include diagnostic category and age category and secondary outcomes will include cost, quality of life, satisfaction, pain, adverse events, rates of imaging, Emergency Department length of stay, treatment time and diagnostic accuracy. Methods of statistical comparison are outlined.

**Conclusion:** This report describes the SAP for the BUCKLED trial, to minimise analysis bias and ensure transparency and internal validity for the findings of this randomised controlled trial.

## Overview

The Bedside Ultrasound Conducted in Kids with distal upper Limb fractures in the Emergency Department (BUCKLED) trial will be the first randomised controlled trial (RCT) comparing point-of-care ultrasound (POCUS) to x-ray as the initial diagnostic imaging modality in non-angulated paediatric distal forearm injuries.(1) We describe the prospectively defined SAP, finalised before patient data is made available for analysis.

## Study Design

The BUCKLED trial is a multicentre, open-label, non-inferiority RCT assessing initial diagnostic imaging modality (POCUS vs x-ray) for paediatric patients with clinically non-angulated distal forearm injuries. Recruitment of up to 300 patients (allowing for attrition) is expected at 4 study sites.

The primary hypothesis is that initial diagnostic imaging with POCUS results in non-inferior upper limb physical function at 4 weeks compared to x-ray, as measured by the Pediatric Upper Extremity Short Patient-Reported Outcomes Measurement Information System (PROMIS) tool. A key secondary hypothesis is the non-inferiority of POCUS compared with x-ray as the initial imaging modality for children who are confirmed to have buckle fractures.

### Patient population

Paediatric patients presenting to a participating Emergency Department (ED) with an isolated, acute, clinically non-angulated, distal forearm injury who are being evaluated for a suspected fracture with imaging.

#### Inclusion Criteria

The inclusion criteria are:

- Age 5 years to 15 years 364 days.
- Distal forearm injury requiring radiological evaluation.
- Ability to attend follow up if required, including living within 50km of attending hospital and having telephone and internet access.

#### Exclusion Criteria

The exclusion criteria are:

- Obvious angulation or deformity (soft tissue swelling allowed).
- Injury sustained > 48 hours prior.
- External x-rays already performed.
- Compound / open fracture.
- Neurovascular compromise.
- Known bone disease (e.g., osteogenesis imperfecta).
- Suspicion of non-accidental injury.
- Additional injuries requiring x-rays (e.g., mid- or proximal forearm, elbow, scaphoid).
- Congenital forearm malformation (e.g., radius hypoplasia)
- No credentialled clinician available to perform scan.
- Significant developmental delay or behavioural difficulties prohibiting accurate clinical assessment.

### Randomisation and Blinding

Randomisation was conducted via a web-based central randomisation service (Griffith University Clinical Trials Randomisation Service) and occurred in a 1:1 ratio within blocks of size 6-8 (randomly selected), stratified by four sites (Gold Coast University Hospital, Queensland Children’s Hospital, Sunshine Coast University Hospital and Robina Hospital) and two age categories (5 – 9 years and 10 – 15 years). The trial is open-label with treating clinicians and participants aware of trial group allocation, as the nature of the imaging modalities studied means that blinding is not possible. However, radiographers performing x-rays and radiology specialists and trainees reporting them will be blinded to group allocation and any POCUS images. The expert panel determining final diagnosis will be blinded to primary and secondary outcome data.

### Intervention

Patients were randomised to undergo either POCUS or x-ray imaging as the initial diagnostic imaging modality. Full details of diagnostic and therapeutic interventions are outlined in the study protocol (1).

#### X-Ray

Patients randomised to x-ray had a minimum of 2 views performed by a radiographer and classified by the treating clinician into a diagnostic category of ‘no’ fracture, ‘buckle’ fracture or ‘other’ fracture. A ‘buckle’ fracture was defined as a deformation of the bony cortex without breach, while ‘other’ fracture was defined as a fracture associated with a cortical breach but also included fractures at an alternative location, such as the scaphoid or proximal two thirds of the forearm. The x-ray group operates as a control comparator and is in keeping with routine ED care.

#### POCUS

Patients randomised to POCUS had imaging performed by a health practitioner with appropriate training and credentialling, consisting of a 6-view forearm POCUS protocol with assessment of secondary signs (1). The resultant images were classified by the treating clinician with an overall forearm diagnosis of ‘no’ fracture, ‘buckle’ fracture or ‘other’ fracture by labelling the final ultrasound image at the conclusion of scanning. When a cortical breach fracture (i.e., ‘other’ fracture) was detected, the patient also received an x-ray. Other pre-specified criteria for x-ray imaging after POCUS included secondary signs of fracture on POCUS or high clinical suspicion, such as for pain out of proportion to clinical findings (see Table 2 of Protocol (1))

#### Management

Patients with ‘no’ fracture were managed at the treating clinician’s discretion. Patients with a ‘buckle’ fracture were managed in a removable wrist splint and patients with an ‘other’ fracture were managed by immobilisation in a plaster cast or equivalent (after manipulation, reduction and/or surgery as required).

### Outcomes

Outcome data was collected at allocation (baseline) and at 1 week, 4 weeks and 8 weeks post-allocation.

#### Primary Outcome

The primary outcome is physical function of the injured upper limb at 4 weeks (28 days ± 3 days), as measured by the PROMIS tool. This tool consists of 8 questions regarding upper limb functions, which are scored from 5, “no trouble” to 1, “not able to do”, such that a score of 40 denotes unimpaired upper limb function and 8 denotes fully impaired upper limb function.

#### Secondary Outcomes

The secondary outcomes are:

- Physical function of the injured upper limb at 1 week (7 days ± 3 days) and 8 weeks (56 days
- ± 3 days) as measured by the PROMIS tool
- Direct and indirect healthcare costs (healthcare provider visits, days off work/school)
- Health related quality of life (QOL), as determined by the Child Health Utility 9D (CHU9D) instrument.
- Satisfaction score (patient and parent)
- Patient pain score measured using Faces Pain Score Revised (FPSR).
- Adverse events (including diagnosis of clinically relevant alternate fracture type and poor fracture healing)
- Rates of x-ray and other imaging (particularly ‘no’ fracture and ‘buckle’ fracture groups)
- ED length of stay (triage to ED discharge)
- Treatment time (clinician review to ED discharge)
- Diagnostic accuracy of POCUS, including secondary signs, and x-ray.

### Sample Size

Sample size calculations were performed to detect a non-inferiority margin of 5 points on the PROMIS tool at 4 weeks, with the estimated standard deviation of the PROMIS tool of 11.5 points (2-4) and estimated between-group difference of 0 points. Using 90% power with one-sided α of 0.025, primary outcome data for 224 patients was required (112 per arm). For the key secondary outcome of non-inferiority of POCUS for patients with a diagnosis of buckle fracture, the estimated standard deviation of the PROMIS tool was 7.5 points (2-4) and estimated between-group difference was 0 points, requiring a total of 96 patients with buckle fractures (48 per arm) to achieve 90% and one-sided α 0.025. The estimated proportion of patients with expert panel diagnosis of buckle fracture was 35 – 45%. To obtain primary outcome data on at least 112 children per arm and at least 48 children per arm with buckle fractures, with a potential withdrawal and loss to follow up rate of 25%, recruitment of up to 300 patients was planned.

### Statistical Analysis

#### Analysis Principles

The analysis principles are:

- All primary analysis will be conducted on an intention-to-treat basis, in which all patients who are randomised are included in the statistical analysis and all patients are analysed in the group to which they are assigned, regardless of expert panel diagnosis, treatment and imaging tests performed. For comparisons with a non-inferiority hypothesis, per-protocol analysis will also be performed and reported alongside intention-to-treat results.
- Statistical significance testing for non-inferiority will be one-sided, with type I error rate (α) of 0.025. Other statistical significance tests will be two-sided with α of 0.05.
- The primary manuscript will include data taken up to 8 weeks (56 days ± 3 days) post randomisation.
- All primary and secondary outcomes of interest will be pre-specified.
- No formal adjustments for multiple testing will be applied. Only analysis findings for the primary outcome will be treated as definitive, whereas secondary outcome findings will be interpreted with due consideration for the multiple comparisons made.
- Categorical variables will be analysed using the χ^2^ test. Alternatively, Fisher’s exact test will be used for tests with small cell sizes (10 or less).
- Continuous variables will be analysed using parametric methods such as linear regression modelling.
- Formal tests of normality will not be performed. For analyses in which marked skew or influential outliers are expected, appropriate non-parametric methods such as median regression modelling will be used.
- Missing data will not be imputed.
- Analysis will be performed primarily using Stata (StataCorp), V14.2 or later.

### Datasets Analysed

The primary analysis will be undertaken on an intention-to-treat basis – patients will be analysed in the group to which they were initially randomised, regardless of the diagnostic tests performed including both protocol-specified crossover and protocol noncompliance. Per-protocol analysis will also be presented for tests with a non-inferiority hypothesis.

### Trial Profile

The flow of patients through the trial will be demonstrated using a flow diagram, consistent with the Consolidated Standards of Reporting Trials (CONSORT) statement (5). This diagram will display the number of patients assessed for eligibility, the number of patients who enrolled and randomised in the study and the number of patients who were excluded or otherwise not enrolled. For enrolled patients, the study group allocation will be displayed, along with actual diagnostic tests received and overall forearm group diagnosis. The number of patients in each group who were lost to follow up or otherwise excluded from analysis will be displayed. Further details are shown in Figure 1.

**Figure 1:**
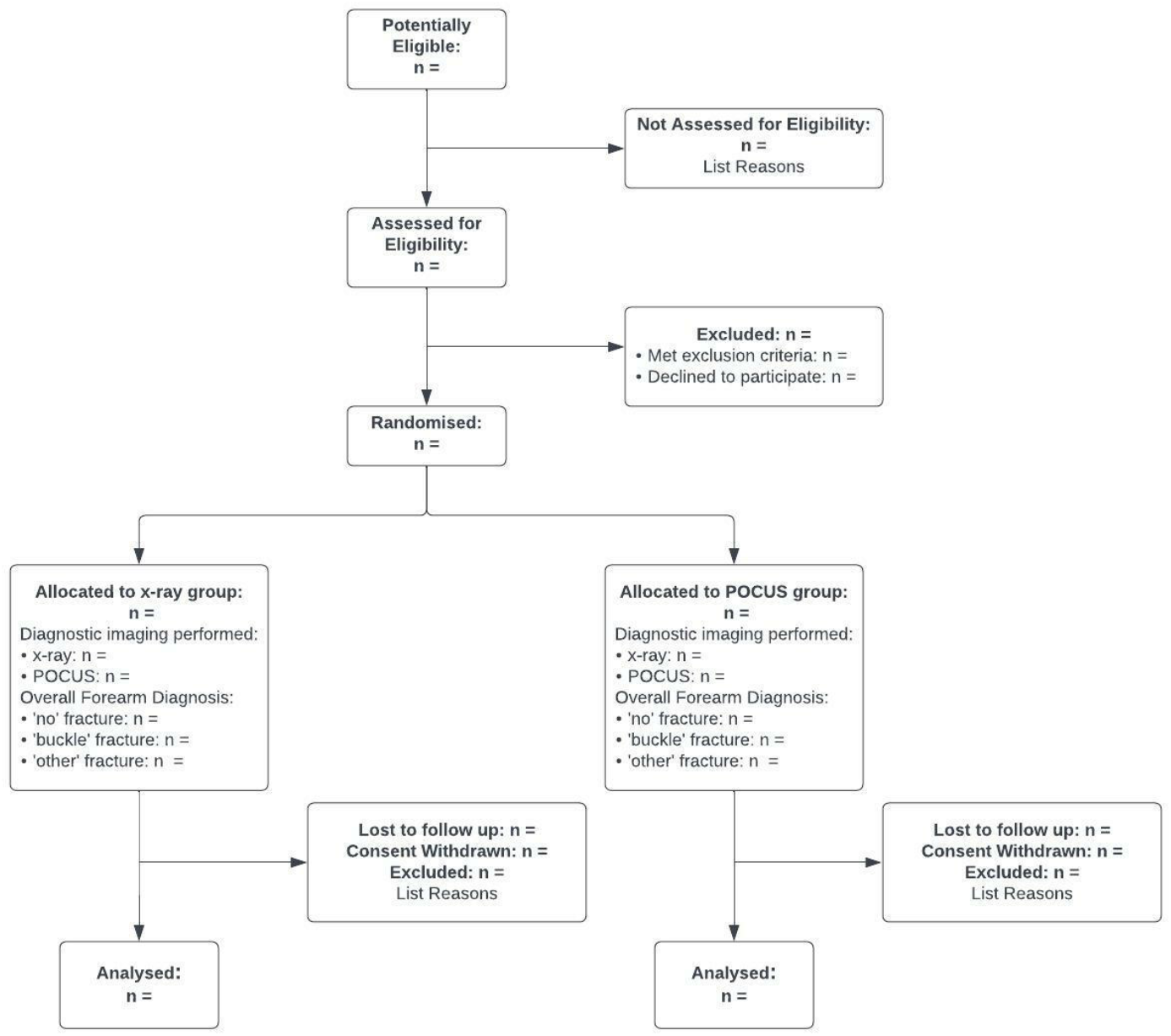
CONSORT Flowchart.

### Patient Characteristics

A description of patient baseline characteristics will be presented by group allocation. Categorical variables will be summarised by frequencies and percentages, using the number of patients with non-missing data as the denominator. Continuous variables will be summarised using either mean with standard deviation (SD) or median with quartiles (25^th^ and 75^th^ percentile) depending on the normality of their distribution.

Baseline characteristics will be tabulated for:

- Sex
- Age described as both a continuous variable and as a categorical variable in keeping with stratification.
- Weight (kg)
- Height (cm)
- BMI percentile for age (6)
- Hand preference
- Affected side
- Previous forearm issue impacting on baseline physical function (injury, surgery, dysfunction)
- Analgesia provided (home and in ED)
- Mechanism of Injury (e.g., fall on outstretched hand, other fall, strike / direct blow, rotational)
- Clinical examination findings (swelling, tenderness, reduced range of motion)

### Overall Forearm Diagnosis and Diagnostic Imaging

Overall forearm diagnosis of ‘no’ fracture, ‘buckle’ fracture or ‘other’ fracture will be tabulated by group allocation, as determined by the treating clinician following initial diagnostic imaging. Patients who receive subsequent diagnostic imaging on the index presentation leading to a change of diagnostic category will be summarised in the text; this will include patients randomised to the POCUS who subsequently do receive x-ray imaging in keeping with the study protocol. The gold standard final diagnosis will be determined by the consensus of an expert panel consisting of a paediatric orthopaedic surgeon, paediatric radiologist, and emergency physician with paediatric emergency fellowship, all otherwise independent to the RCT, who will consider the clinical course, investigation findings and management.

Diagnostic studies performed by treatment allocation will be summarised on the CONSORT diagram and described in the text. For patients who are randomised to the POCUS group and subsequently receive x-ray imaging as part of the index ED presentation, the clinical indications for x-ray imaging will be tabulated and may include:

- Cortical breach fracture identified
- Buckle fracture < 1 cm from physis
- Pronator quadratus haematoma present (7)
- Angulation greater than ∼ 5 degrees
- Physis widened or narrowed
- Periosteal haematoma
- Clinical suspicion e.g., pain out of proportion to ultrasound findings.
- Indeterminate or inadequate ultrasound images
- No reason given (i.e., protocol violation)

### Emergency Department Management

A summary of emergency department management will be presented by group allocation. Categorical variables will be summarised by frequencies and percentages, using the number of patients with non-missing data as the denominator. Continuous variables in this section are expected to have a significant positive skew and be summarised using median with quartiles (25^th^ and 75^th^ percentile).

Characteristics of ED management will be tabulated, including:

- Analgesia received
- Placement of bandage, splint, or cast (above or below elbow; backslab or complete)
- Requirement for manipulation in ED (moulding, traction / reduction), manipulation (traction / reduction) in the operating theatre or surgical fixation.
- Time to review (triage to clinical review)
- Time to imaging (triage to imaging initiation)
- ED length of stay (triage to ED discharge)
- Treatment time (clinician review to ED discharge)

Duration of bandage, splint or cast placement and number and type of follow up reviews (ED, orthopaedic outpatient clinic, general practitioner, and allied health) over the follow up period will also be reported.

### Primary Outcome

#### Main Analysis

The primary outcome is upper limb function as measured by the PROMIS tool at 4 weeks following randomisation. The between-group difference in upper limb function at 4 weeks will be assessed using linear regression modelling with trial group allocation (POCUS / x-ray) included as a main effect. Using the pre-specified non-inferiority margin of 5 points on the PROMIS scale, non-inferiority will be established if the 95% confidence interval (CI) for the effect of allocation to the POCUS group compared to the x-ray group is above this margin. Between-group mean difference and 95% CI for the effect of trial group allocation will also be reported. Missing data will not be imputed.

#### Subgroup Analysis

A pre-specified key secondary analysis is the non-inferiority of upper limb function at 4 weeks by treatment group for patients who are determined to have ‘buckle’ fractures on final diagnosis following expert panel review, as well as for patients determined to have ‘buckle’ fractures by treating clinician diagnosis. For both cohorts, the difference in upper limb function by trial group allocation (POCUS / x-ray) will be assessed with linear regression modelling with a pre-specified non-inferiority margin of 7.5 points on the PROMIS scale. Other subgroup analyses will include each final diagnostic category (‘no’ fracture and ‘other’ fracture, in addition to ‘buckle’ fractures as above) and age category (5-9 years and 10-15 years). Results of subgroup analysis will be displayed on a Forest plot.

### Analysis of Secondary Outcomes

Analysis of secondary outcomes will include direct and indirect health care costs, health related quality of life using CHU9D, patient and parent satisfaction, pain, adverse events, rates of imaging, ED length of stay and treatment time. Main analysis of the secondary outcomes of health-related quality of life, patient and parent satisfaction and pain, as well as the primary outcome of upper limb function, will be performed using data collected at 4 weeks following randomisation. Additional analyses of primary and secondary outcome data collected at 1 week and 8 weeks will also be performed. Proposed analyses are summarised in Table 1.

**Table 1:**
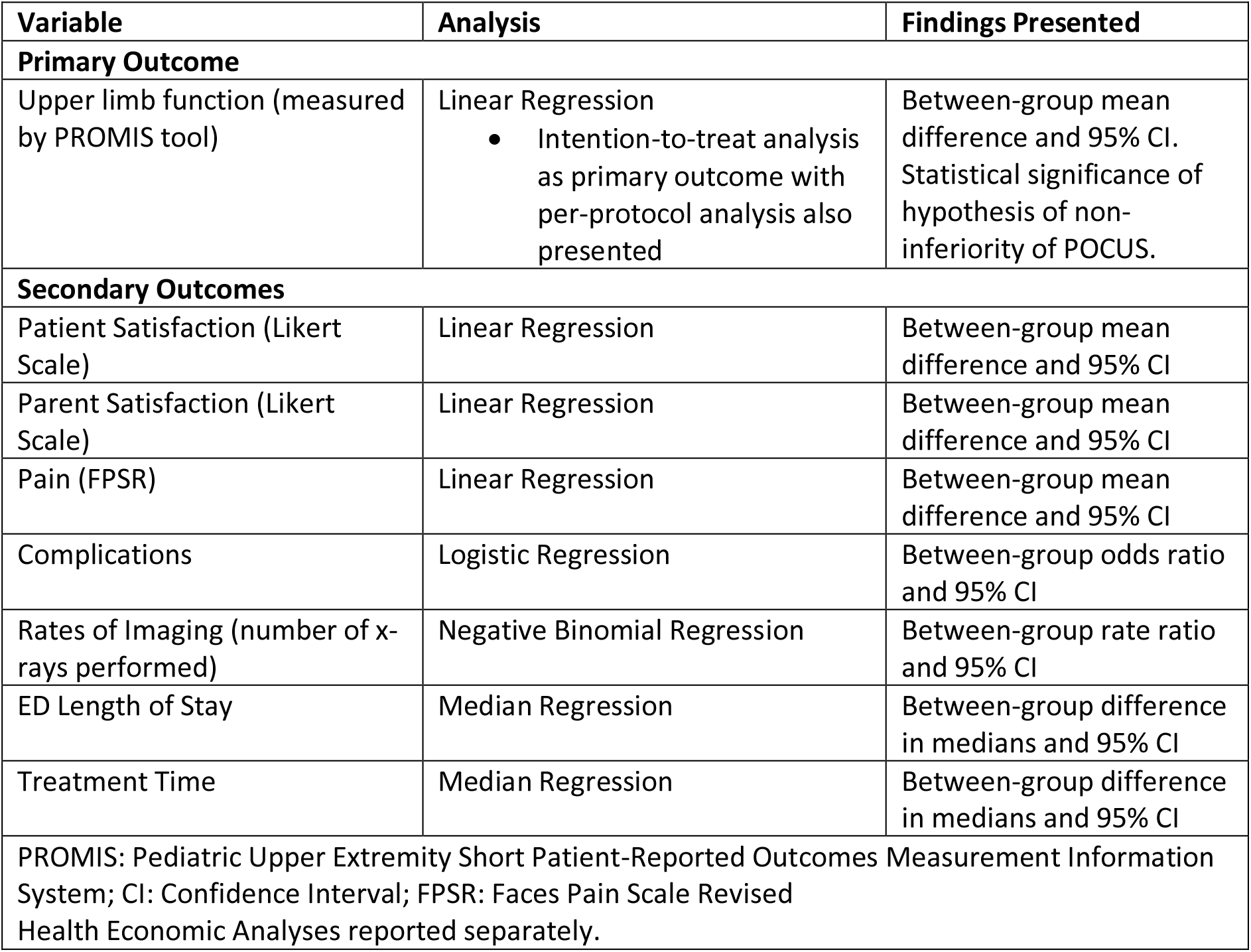
Outcome Measures and Planned Analysis.

#### Health Economic Analysis

The primary economic analysis is a cost-utility analysis from the healthcare provider perspective. The cost-utility analysis will be conducted on a complete case basis with two summary outcome measures, the incremental cost-utility ratio (ICUR) and the incremental net monetary benefit (INMB). The ICUR is defined as Δ*C*/Δ*QALY*; where Δ*C* is the incremental cost per patient and Δ*QALY* is the incremental quality adjusted life years per patient, with a treatment option considered cost-effective where the ICUR is less than λ, the pre-defined willingness-to-pay threshold of a QALY. The INMB is defined as Δ*QALY* × *λ* − Δ*C* and a treatment option considered cost-effective where INMB is < 0. A priori, the value of λ is considered $50,000 per QALY; an implied threshold commonly used within the Australian setting (8).

For each individual, QALYs will be estimated using the Area Under the Curve (AUC) approach, in which the change in health-related quality of life utility scores is assumed to occur linearly between measurements. The unadjusted difference in QALYs between the randomisation groups will be calculated as per Eq (1).

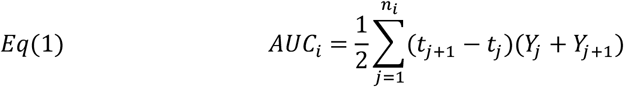

Where *Y*_*ij*_ represents the health-related quality of life utility score observed at time *t*_*ij*_, for observation j = 1, …, *ni* on subject *i* = 1, …, *m*.

Health related quality of life utility scores will be derived from the CHU9D scores using the Australian adolescent-specific scoring algorithm. (9) Secondary analysis of change in health related quality of life utility values between baseline and follow up will be undertaken using multi-level regression modelling with a random intercept, including time as a repeated categorical variable and adjusting for baseline scores (10).

From a healthcare system perspective, costs included in the analysis will be the sum of all directly related healthcare costs and include costs associated with both the initial delivery of care as well as any subsequent healthcare costs attributable to initial procedures. The mean and standard deviation of the direct healthcare costs for both groups will be presented separately for costs associated with the initial treatment procedures, subsequent healthcare utilisation and total direct healthcare costs. As costs are usually non-normally distributed (11), we will use generalised linear regression models (a priori assuming log link and gamma error, which will be tested during analysis using the Pregibon link test and modified Park’s test) to estimate the mean difference in costs between groups.

Uncertainty in the ICUR and INMB estimates will be characterised using bootstrapping (minimum 1,000 iterations) with 95% credible intervals being reported. Uncertainty in the ICUR will also be visually depicted using a cost-effectiveness plot. Sensitivity to accepting the new intervention as cost effective to the threshold value of a quality adjusted life year (λ) will be assessed using a cost-effectiveness acceptability analysis (12).

Subsequent scenario analyses will be undertaken from a broader societal perspective including the direct and indirect costs borne by patients and their caregivers with respect to time and travel costs. These will be estimated using the Cost & Management survey which has been adapted for use in this study (13).

#### Patient and Parent Satisfaction

Satisfaction scores were collected independently from patients and parents, based upon a 5-point Likert scale (14). Analysis of Likert scale scores collected at 4 weeks will be used to compare satisfaction in the POCUS and x-ray groups for both parents and children using linear regression modelling. Despite the ordinality of Likert scale data, it is well established that parametric statistical methods such as linear regression are appropriate for analysing data of this form, as these methods are robust to significant non-normality in the underlying distribution (15). Effect estimates will be presented as between-group mean difference and 95% CI.

#### Pain

Pain scores were collected using the Faces Pain Score Revised. Analysis of FPSR scores collected at 4 weeks will be used to compare pain levels between the POCUS and x-ray groups. Linear regression modelling will be performed. Effect estimates will be presented as between-group mean difference and 95% CI.

#### Adverse Events

Adverse events will be compared across the two study groups, including diagnosis of a clinically relevant alternative fracture pattern to initial imaging, worsening deformity, rates of re-injury, growth disturbance, delayed fracture healing and/or requirement for surgical intervention. The number of patients who sustain at least one adverse event will be reported for each trial group. Proportions of patients with at least one adverse event will be compared between the study groups using logistic regression with group allocation as a main effect. Effect estimates will be presented as between group odds ratio and 95% CI

#### Rates of Imaging

Rates of imaging required will be compared between the trial arms. The proportion of patients requiring advanced imaging, such as CT or MRI, will be compared using the χ^2^ test for each imaging modality. The total number of x-rays performed will be compared between study groups using Poisson regression modelling with group allocation as a main effect. Alternatively, negative binomial regression will be used if Poisson regression demonstrates evidence of lack of fit associated with overdispersion. A prespecified subgroup analysis will be total number of x-rays performed by trial allocation, in patients diagnosed with ‘no’ or ‘buckle’ fractures on their index presentation. This comparison will be tested using Poisson or negative binomial regression modelling as above. Effect estimates will be presented as rate ratios with 95% CI.

#### ED Length of Stay and Treatment Time

The ED length of stay and treatment time will be determined from the electronic clinical record. The distributions of these variables are expected to demonstrate significant positive skew, and so the ED length of stay and treatment time for the POCUS and x-ray groups will be summarised using median and interquartile range, with median regression used to evaluate statistical significance. Effect estimates will be presented as between-group difference in medians and 95% CI.

### Additional Studies using BUCKLED RCT Data

Two key additional studies will use BUCKLED RCT data to evaluate (i) the accuracy of POCUS, compared to X-ray, for diagnosing upper arm fractures, and (ii) the diagnostic value of secondary signs measured using POCUS to determine upper arm fracture. The results of each of these studies will be reported in separate manuscripts.

#### Diagnostic Accuracy of POCUS and X-ray

The diagnostic accuracy of POCUS and x-ray imaging on index presentation will be evaluated against the final diagnosis as determined by the expert panel. The analysed data set will consist of all BUCKLED RCT participants who received a clinical diagnosis based on either POCUS or an X-ray and whose images and clinical history were assessed by the expert panel. A case is defined as ‘true positive’ when both the initial diagnostic imaging modality (POCUS or x-ray) and the expert panel diagnose any fracture type (i.e., ‘buckle’ fracture and/or ‘other’ fracture). A case is defined as ‘true negative’ when the initial diagnostic imaging modality and expert panel both diagnose ‘no’ fracture. Given the increased clinical significance of ‘other’ fractures, a secondary analysis of diagnostic accuracy will combine the ‘no’ fracture and ‘buckle’ fracture groups, such that a case is defined as ‘true positive’ when both POCUS or x-ray and the expert panel diagnose an ‘other’ fracture, ‘true negative’ when the initial diagnostic imaging modality and the expert panel diagnose any combination of ‘no’ or buckle’ fracture, and so forth. Sensitivity and specificity will be tabulated for the POCUS and x-ray imaging groups. 95% confidence intervals will be calculated using the Wilson method for calculating the confidence interval of a proportion. The statistical significance of the difference in sensitivity and specificity will be calculated using a normal approximation of the difference in proportions.

#### Diagnostic Accuracy of POCUS Secondary Signs

The ability of secondary signs measured during POCUS to diagnose fracture type will be investigated. Participants will contribute data to this study if they received a clinical diagnosis after receiving POCUS and had an expert panel assessment. Secondary signs investigated will include the distance of the fracture from the physis and the presence of the pronator quadratus haematoma sign. Each secondary sign investigated will be summarised according to the three fracture types. Receiver operating characteristic curves will be generated for each of the secondary signs, to identify appropriate cut-points. Diagnostic statistics will then be calculated, and the sensitivity and specificity compared.

## Data Availability

All datasets used and/or analysed during the BUCKLED RCT will be available from the corresponding author of that paper upon reasonable request and after requestees meet criteria stipulated by the approving ethics committee.

## Proposed Figures

Figure 1. CONSORT Flowchart

Figure 2. Difference in PROMIS Score at 4 weeks by Group Allocation

Figure 3. PROMIS Score Subgroup Analysis Forest Plot

## Proposed Tables

Table 1. Baseline Characteristics by Group Allocation

Table 2. Overall Diagnostic Category by Initial Imaging Modality, Index Presentation and Expert Panel Diagnosis

Table 3. Indications to Perform X-ray Imaging in Patients Randomised to POCUS

Table 4. ED Management Characteristics by Group Allocation

Table 5. PROMIS Score by Group Allocation

Table 6. PROMIS Score Subgroup Analysis

Table 7. Satisfaction and Pain Scores

Table 8. Complications, Rates of Imaging, ED Length of Stay and Treatment Time.

## Declarations

### Authors’ Contributions

This statistical analysis plan was written by PMJ, with input and advice from PJS, RSW, GK and JB. PJS conceived and designed the BUCKLED RCT. RSW, GK and PMJ advised on the statistics, including sample size and analysis plan. JB advised on the health economics. All authors read and approved the final manuscript. PMJ takes responsibility for the contents of this SAP.

### Ethical Approval and Consent to Participate

Multisite ethical approval for the trial was granted by Children’s Health Queensland Hospital and Health Service Human Research Ethics Committee (HREC/19/QCHQ/53306). Informed consent was obtained from all trial participants. This trial was prospectively registered at ANZCTR (ACTRN12620000637943)

